# Should the governance of individual treatment attempts (“Individuelle Heilversuche”) include praxis evaluation? Results from qualitative stakeholder interviews

**DOI:** 10.1101/2022.03.21.22272689

**Authors:** Alice Faust, Lena Woydack, Daniel Strech

## Abstract

**Introduction:** Individual treatment attempts (ITAs) are a German concept for the treatment of individual patients by physicians with nonstandard therapeutic approaches. ITAs span from nonstandard off-label drug uses to first-in-human uses of newly developed drugs/ interventions. Due to the lack of evidence, ITAs come with a high amount of uncertainty regarding the risk-benefit ratio. At present, no prospective review and no systematic retrospective evaluation of ITAs are required in Germany; therefore, no opportunity exists for the basic evaluation of the frequency, type, or outcomes of ITAs. Our objective was to explore stakeholders’ attitudes toward the retrospective evaluation (monitoring) or prospective evaluation (review) of ITAs.

**Methods:** We conducted a qualitative interview study among relevant stakeholder groups. We used the SWOT framework to represent the stakeholders’ attitudes. We applied content analysis to the recorded and transcribed interviews in MAXQDA.

**Results:** Twenty interviewees participated in the study. The interviewees pointed to several arguments in favor of the retrospective evaluation of ITAs, such as the knowledge gain about and setting and circumstances of ITAs. At the same time, the interviewees expressed concerns regarding the validity and practical relevance of the evaluation results. The viewpoints on review addressed several contextual factors (e.g., different medical disciplines) that should be acknowledged when judging the necessity of a review.

**Conclusion:** One main conclusion is that the current situation with a complete lack of evaluation insufficiently reflects safety concerns. German health policy decision makers should be more explicit about where and why some sort of evaluation is needed or not needed. Another conclusion is that there is no one-size-fits-all model for evaluating ITAs. Both prospective and retrospective evaluations should be piloted in areas of ITAs with particularly high risks and conflicts of interest, such as in the application of very experimental therapies (e.g., bench-to-bedside applications) outside clinical trials.

---

What is already known on this topic? In Germany, the concept of individual treatment attempts (ITAs) or “Individuelle Heilversuche” exists for the treatment of patients when standard therapies have failed. At present, no systematic evaluation of the practice, setting and circumstances of ITAs exists.

What this study adds? This is the first study to qualitatively investigate stakeholder viewpoints for and against the retrospective or prospective evaluation of ITAs.

How might this study affect research, practice or policy? This study is intended to raise awareness of the lack of data and guidelines that are needed for the responsible governance of medical care affected by high uncertainty, risks, and conflicts of interest. The study results further provide a reference to guide future discussion and policy development on whether, how and where ITAs should be accompanied by evaluation.

## Introduction

In 2015, a seven-year-old child suffering from junctional epidermolysis bullosa (JEB) was admitted to the Children’s Hospital in Bochum (Germany) [1]. The child’s condition worsened because of severe bacterial infections, and he eventually experienced an epidermal loss of 80%. All therapeutic options had failed [1]. In such desperate situations, physicians in Germany can conduct individual treatment attempts (ITAs, in German: “Individuelle Heilversuche”). In the case of the boy, the physicians transplanted his skin regenerated with transgenic epidermal stem cells. This procedure was highly experimental and had only been used twice in other humans before. No clinical trial had previously investigated this intervention, and no trial was ongoing at the time.

ITAs are a German concept for the treatment of individual patients by physicians with nonstandard therapeutic approaches [2]. The German Federal Supreme Court defines the medical standard as follows: “The respective state of scientific knowledge and medical experience that is necessary to achieve the medical treatment goal and has proven itself in testing” [3]. Scholars ethically justify ITAs as an ultima ratio method that is deployed when all therapeutic options have failed or there is no standard therapy available [4, 5]. The term nonstandard therapy is very broad, and thus, ITAs span from nonstandard off-label drug uses to first-in-human uses of newly developed interventions.

As nonstandard therapies often lack evidence from clinical trials, ITAs are associated with increased uncertainty regarding their risk-benefit ratio [2]. This uncertainty addresses several ethical challenges [6] that have also been discussed in the context of experimental or unapproved therapies [7-9], off-label uses [10, 11], or first-in human therapies [12]. For example, the higher the uncertainty is, the more intuition-based the assessment of risks and benefits. Intuitions are susceptible to different kinds of bias and can thus increase the likelihood of misjudgment about risks and benefits [13].

In other contexts of medical care that are prone to high levels of uncertainty (e.g. organ transplantations, care of premature infants) or in medical research, the actual effects of practices are often evaluated retrospectively based on systematic documentation [14, 15]. International guidelines such as the Declaration of Helsinki (paragraph 37) recommend documentation and evaluation as basic ethical principles for treatment attempts with individual patients with high uncertainty [16]. For ITAs in Germany, no form of systematic documentation and thus no retrospective evaluation for ITAs exist. At present, it is impossible to know what and how many ITAs have been conducted in Germany. As a consequence, no evaluation exists about how often ITAs (in a certain therapeutic area) result in substantial health benefits and/or severe side effects.

Furthermore, guidelines, e.g., from university medical centers and professional societies in the USA, suggest a form of prospective review of innovative care attempts to ensure patients’ safety [17, 18]. While several ITAs in the German context might be classified as innovative care, no standardized process for an independent review exists in Germany. The possibility of contacting a clinical ethics expert for physicians who want to conduct ITAs is mentioned on the web pages of a few university medical centers [19, 20].

We conducted a qualitative interview study to explore stakeholders’ attitudes toward retrospective evaluation (monitoring) and prospective evaluation (review) of ITAs. We categorized these attitudes in terms of strengths, weaknesses, opportunities and threats (SWOTs). Furthermore, we gathered stakeholders’ suggestions for different options regarding retrospective or prospective evaluation.

## Methods

### Ethics approval

The research ethics committee of Charité Universitätsmedizin Berlin approved our study (EA4/005/21).

### Sampling

We used a purposive sampling strategy combined with a snowballing approach to identify potential participants with different backgrounds relevant to the topic of ITAs (physicians, bioethicists, a legal expert, a health care insurance representative, patient representatives, and representatives of the level of self-administration in the German health care system). The initial sampling strategy included searches across webpages of different hospitals and stakeholder groups as well as the professional networks of DS and LW. Invitations (*supplementary 1*) together with a consent form (*supplementary 2*), study information (supplem*entary 2*) and a data protection declaration (*supplementary 2*) were sent out via e-mail.

### Procedure

We performed qualitative in-depth interviews [21] that lasted between 30-50 minutes between March and November 2021. We used audio-recorded video or phone calls for the interviews and took field notes during the interviews. The audio recordings were transcribed by the company Amberscript under a data processing agreement. The participants did not provide feedback on the findings. A physician (AF) with experience in qualitative research conducted all interviews. Both LW and DS, both physicians and experienced qualitative researchers, were present during 5 (LW) and 5 (DS) interviews to supervise and support AF. We developed a first interview guide with open questions for the semistructured interviews [22]. We slightly adapted the initial interview guide after conducting 10 interviews with mainly clinicians and some ethicists. In the first version of the guide (*see supplementary 3*), we asked about different aspects of a retrospective evaluation (e.g., documentation or data use) separately. We adapted the interview guide (*see supplementary 4*) because the interviewees did not address the different aspects of retrospective evaluation (e.g., documentation) separately, and a slightly adapted procedure was necessary for interviews with stakeholders from nonclinical areas.

### Data analysis & definitions

AF analyzed the transcripts with MAXQDA using content analysis [23]. LW read all the transcripts and double-coded difficult passages. AF and LW discussed the coding results. Only minor discrepancies occurred, and they could all be resolved through discussion. We discussed the codebook and the results in the whole team (AF, LW, DS) multiple times until everyone agreed on the final codebook. We employed deductive and inductive category formation [24]. The deductive categories “retrospective evaluation/monitoring” and “prospective evaluation/review” were extracted from the interview guide. As first-order subcategories, we used “SWOTs” and “Strategies” for retrospective and prospective evaluation. Second-order and further subcategories were derived in an inductive manner directly from the material. From interviews with physicians, we obtained additional results on subtypes of ITAs. We summarized these results under the theme “Categories of ITAs.” Thematic saturation was reached at interview 17, when no new themes could be identified [25].

## Results

We invited 56 people to participate, of whom 20 from different professional areas participated. The physicians had varying degrees of professional experience, and all worked at university medical centers. See table 2 for the participants’ characteristics.

**Table 1:** Our definition of SWOTs is based on a definition employed by Wieschowski et al. [26]

|  |  |
| --- | --- |
| Strengths | The inherent qualities of prospective and retrospective evaluation that have a positive impact on patients, the health care system, or the public in general. |
| Weaknesses | The inherent qualities of prospective and retrospective evaluation that are harmful for patients or the health care system, or the public in general. |
| Opportunities | The external and procedural factors that could support an implementation of a prospective and retrospective evaluation of ITAs. |
| Threats | The external and procedural factors that could threaten an implementation of a prospective and retrospective evaluation of ITAs. |

What we classify as SWOTs in each case reflects the opinions of the interviewees and not necessarily the authors' opinions.

**Table 2:** Participant characteristics

|  |  |
| --- | --- |
| **Total** | 20* |
| **Gender** |  |
| Male | 14 |
| Female | 6 |
| **Stakeholder group** |  |
| Physician | 12 |
| Bioethicist | 3 |
| Legal expert | 1 |
| Health care insurance representative | 1 |
| Representative of the level of self-administration in the German health care system | 3 |
| Patient representative | 2 |
| **Clinical area (of** |  |
| **physicians)** | 5 |
| Oncology | 1 |
| Translational oncology | 2 |
| Nuclear medicine | 1 |
| Intensive care medicine | 2 |
| Pediatrics | 2 |
| Neurology |  |
| **Region (of workplace)** |  |
| Southern Germany | 5 |
| Western Germany | 4 |
| Eastern Germany (with Berlin) | 6 |
| Northern Germany | 5 |

\*The participants could belong to more than one stakeholder group and be trained in more than one clinical area

We structured our results under the following three themes: “Retrospective Evaluation (Monitoring)”, “Prospective evaluation (Review)” and “Categories of ITAs”. In the following, we present the key points of our findings that were discussed most intensively or extensively by the interviewees in a narrative form. For the spectrum of all our identified themes, see Tables 3 (E1-45, R1-15) and 4 (S1-S3), which give example quotes for every identified code.

**Table 3*:**
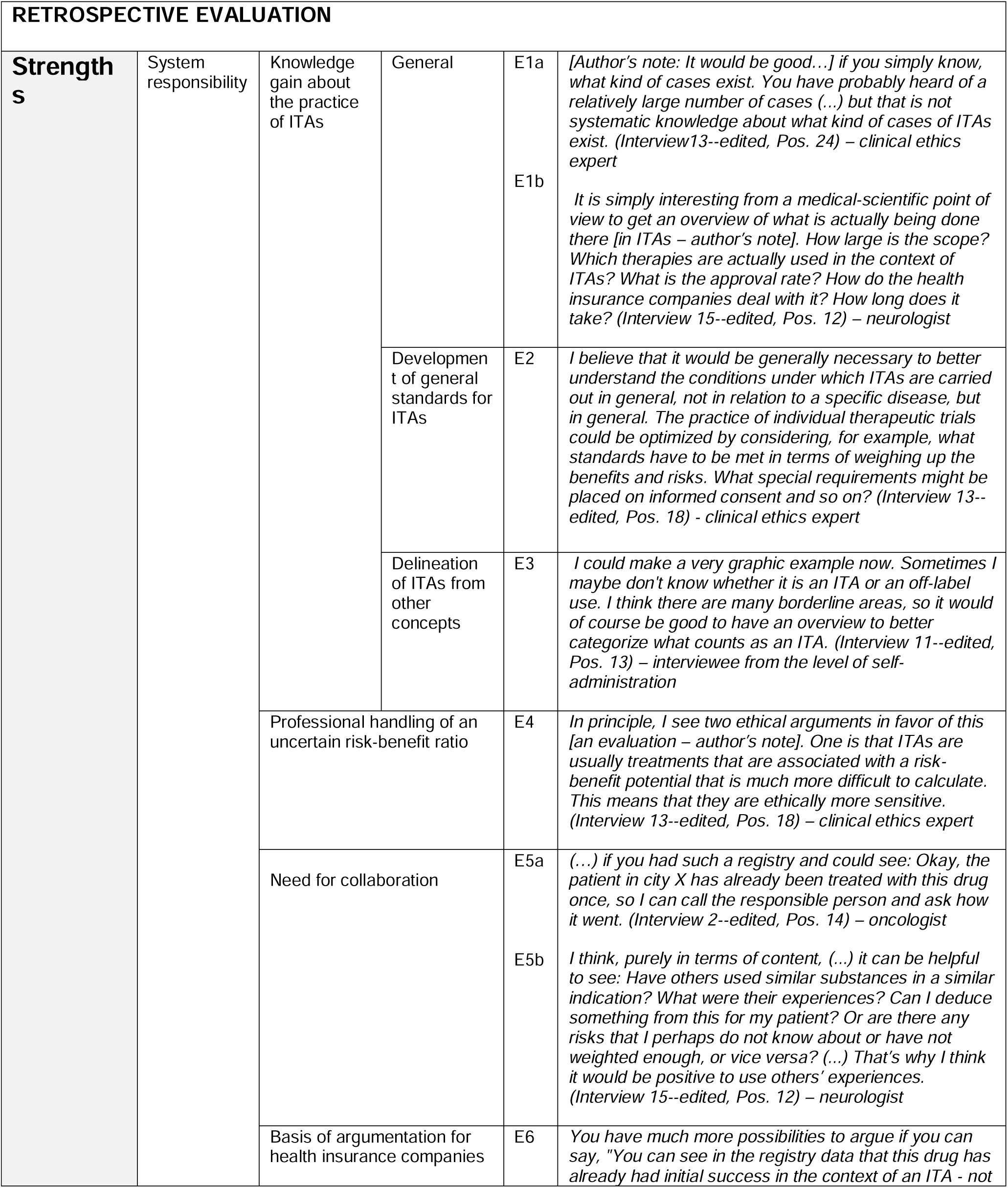
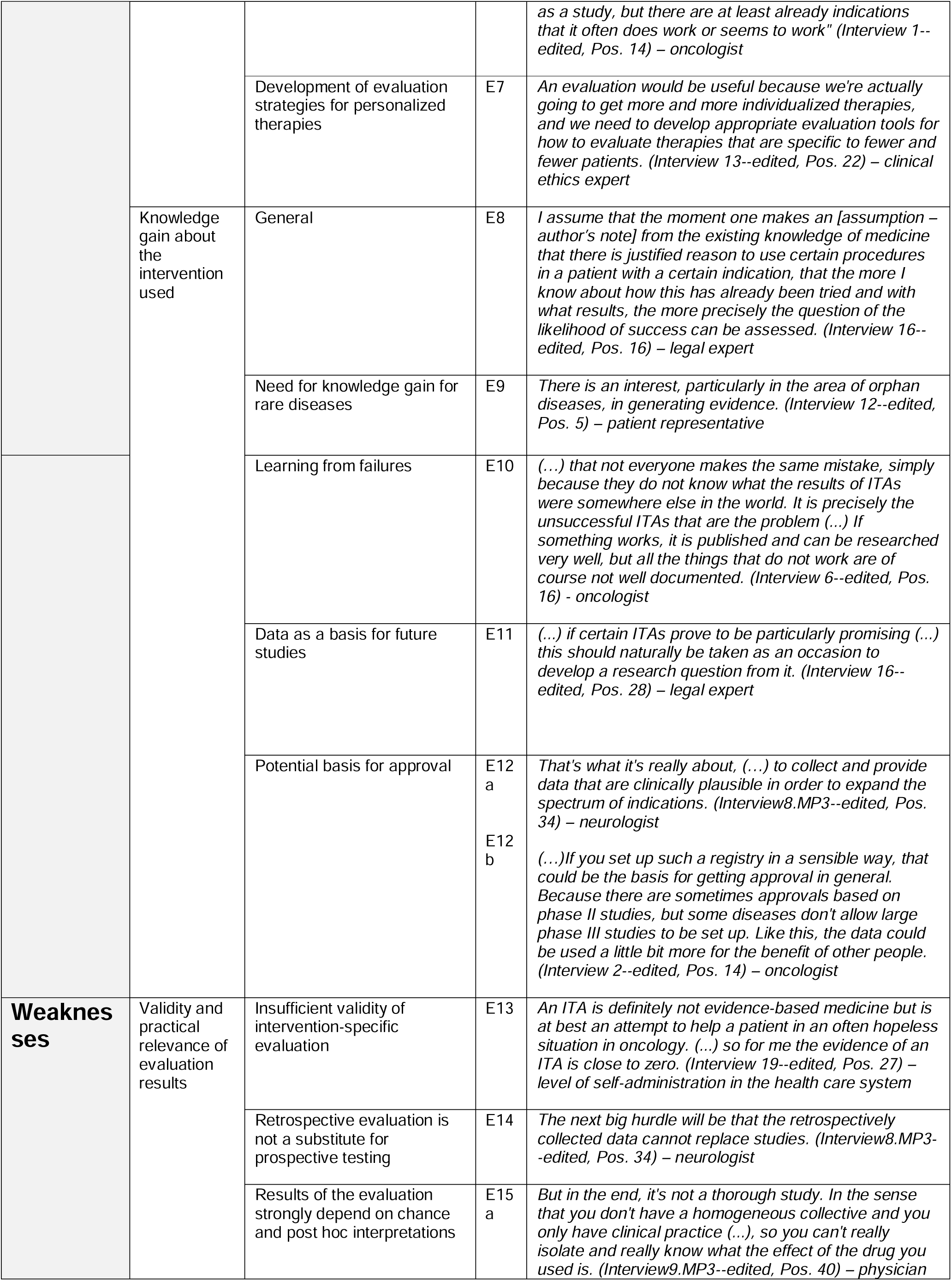
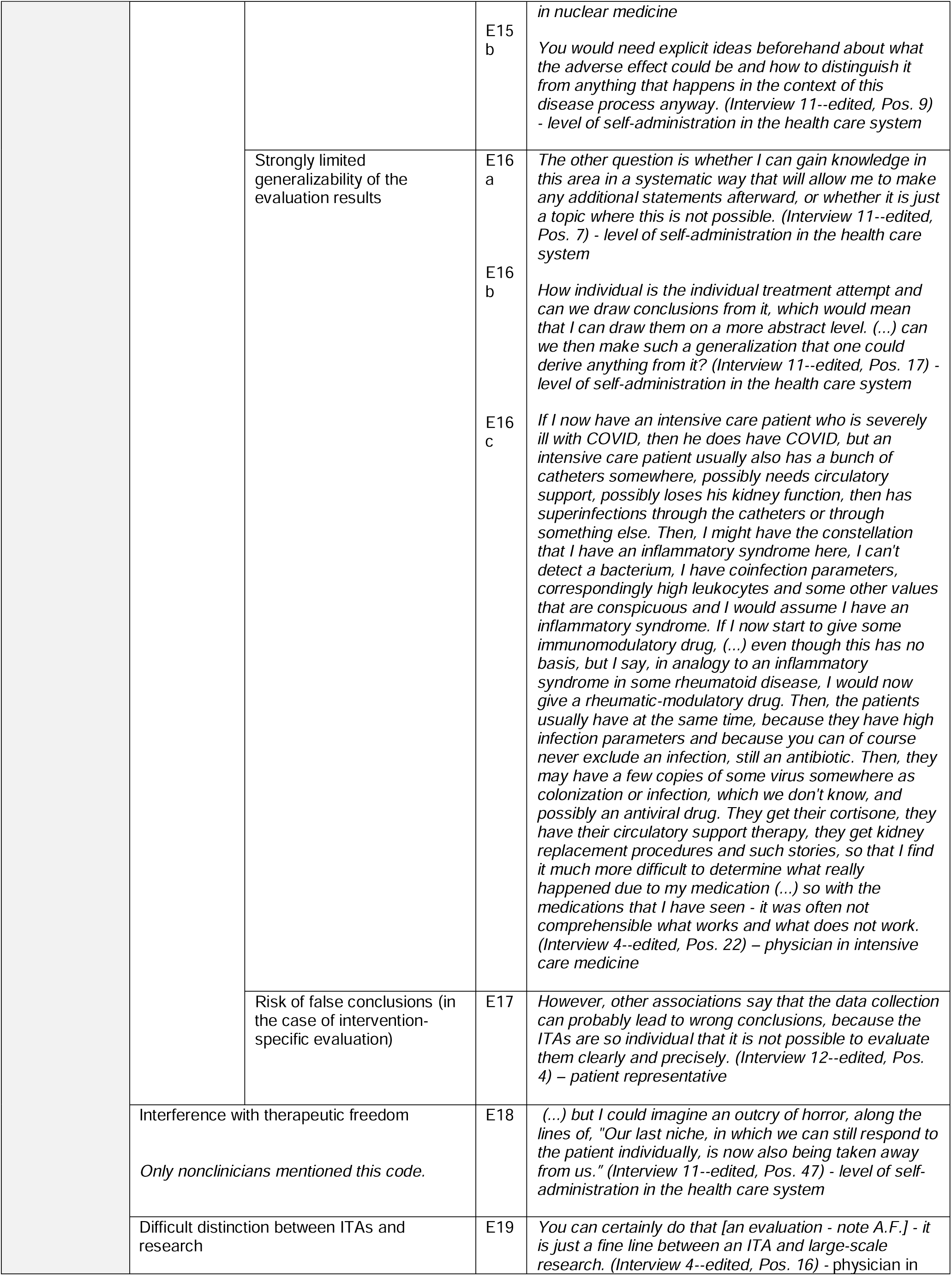
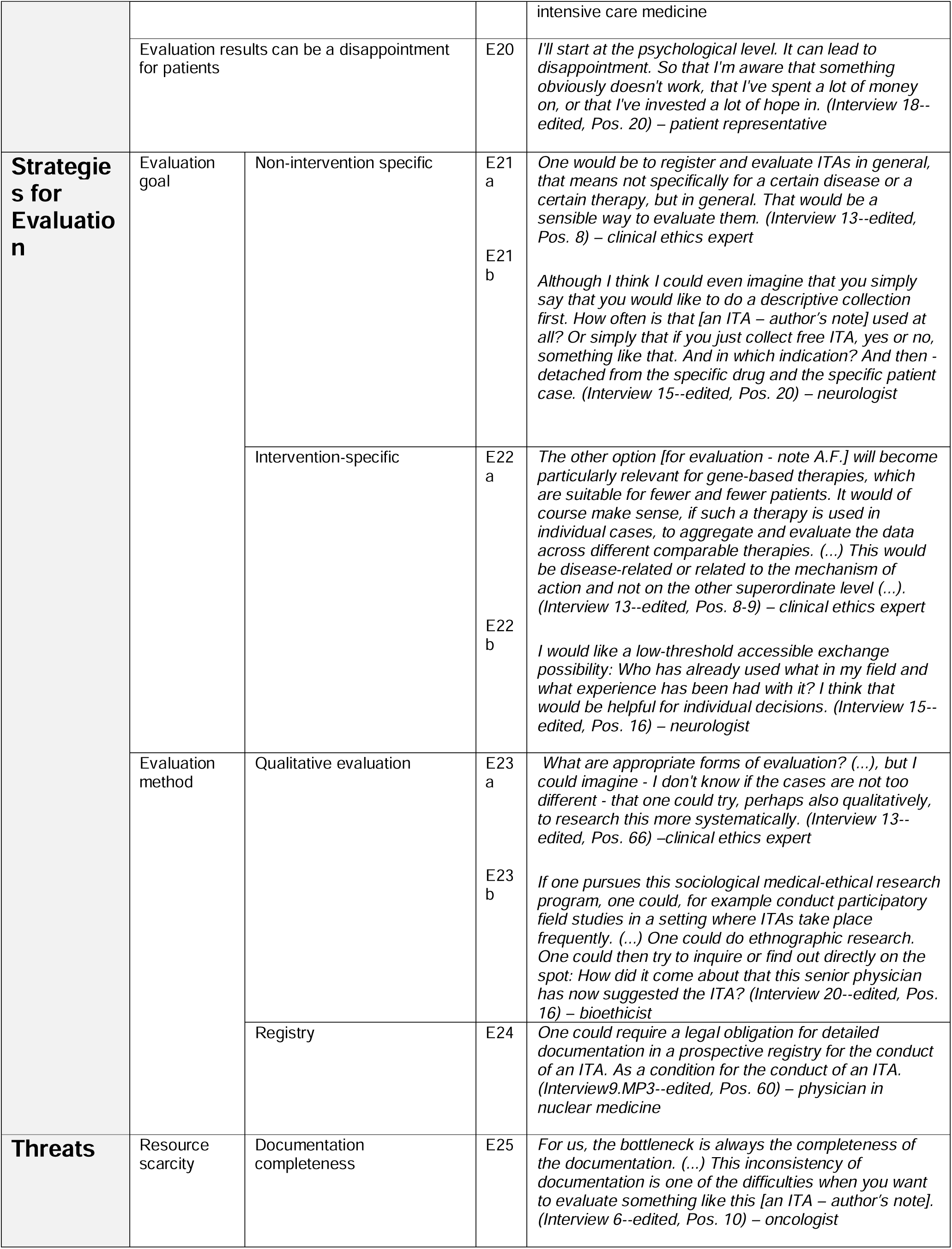
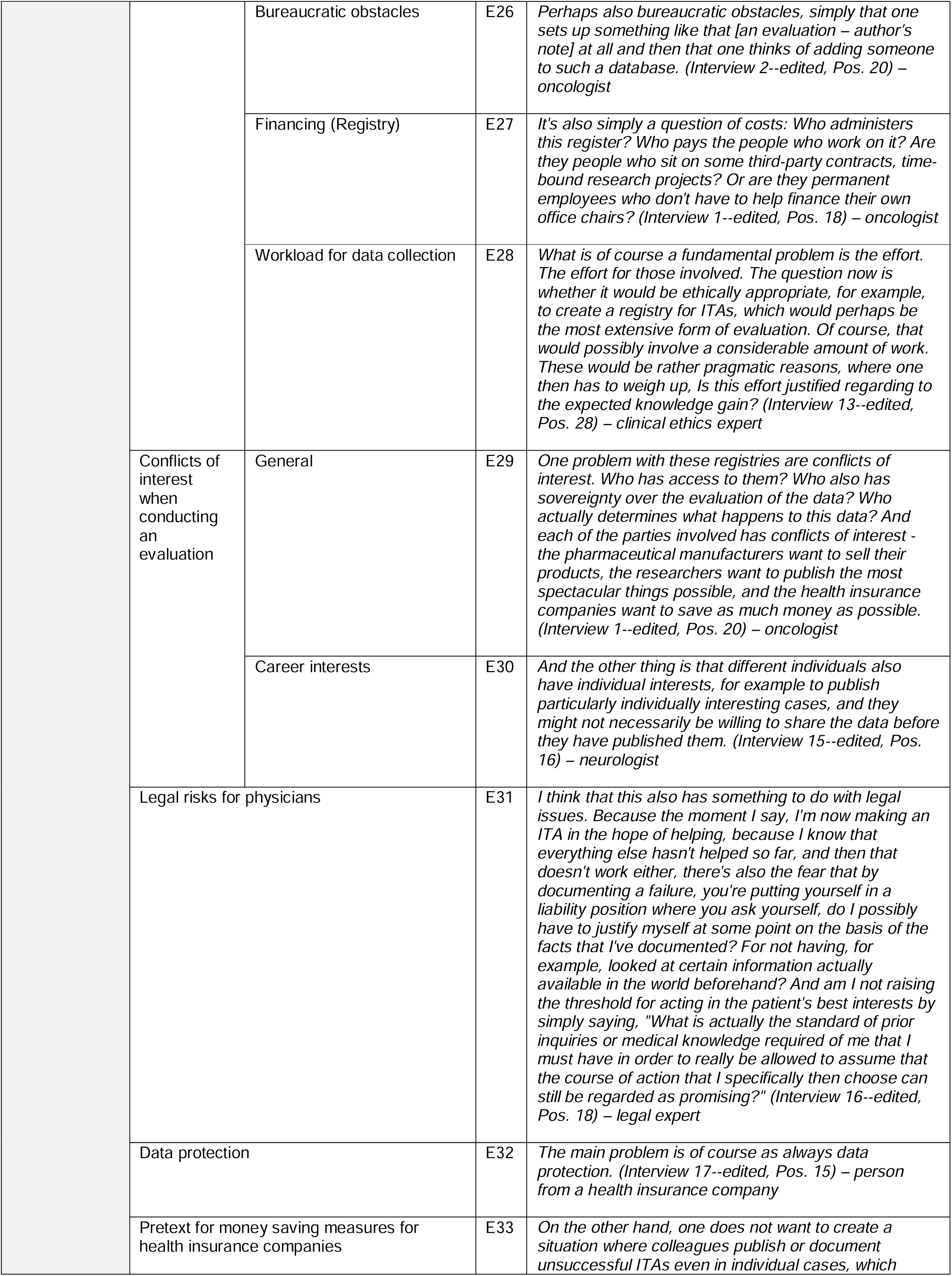
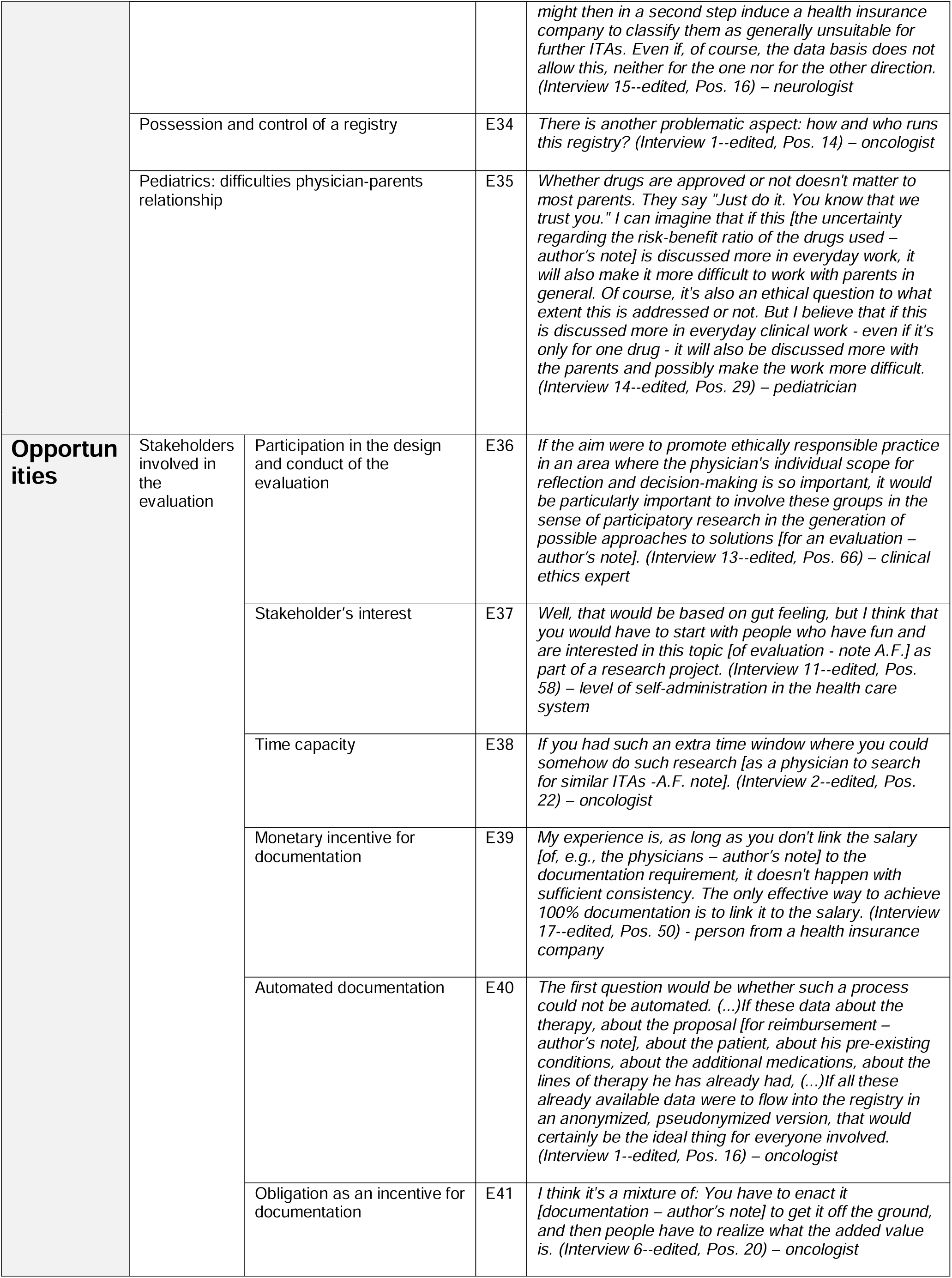
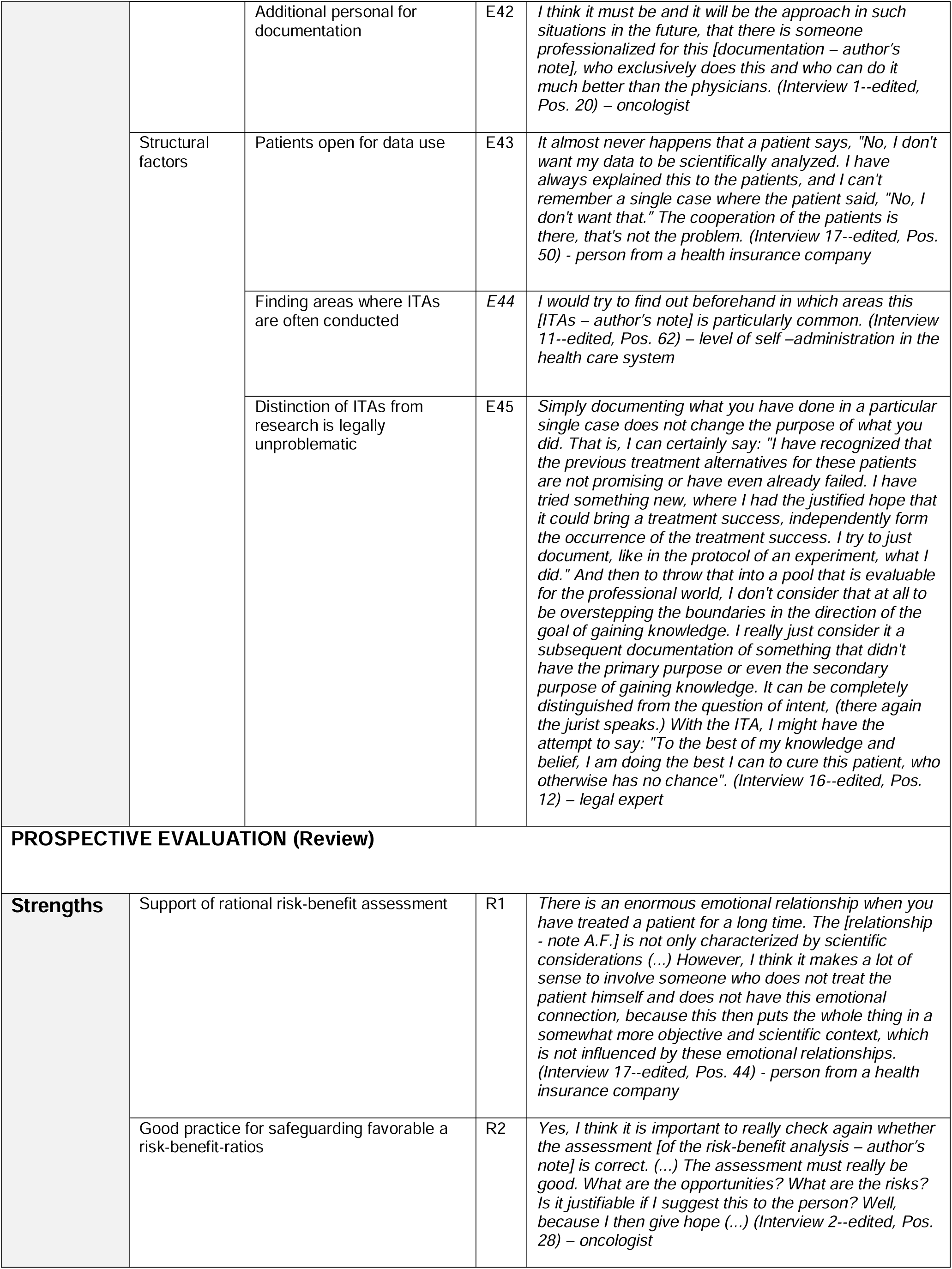
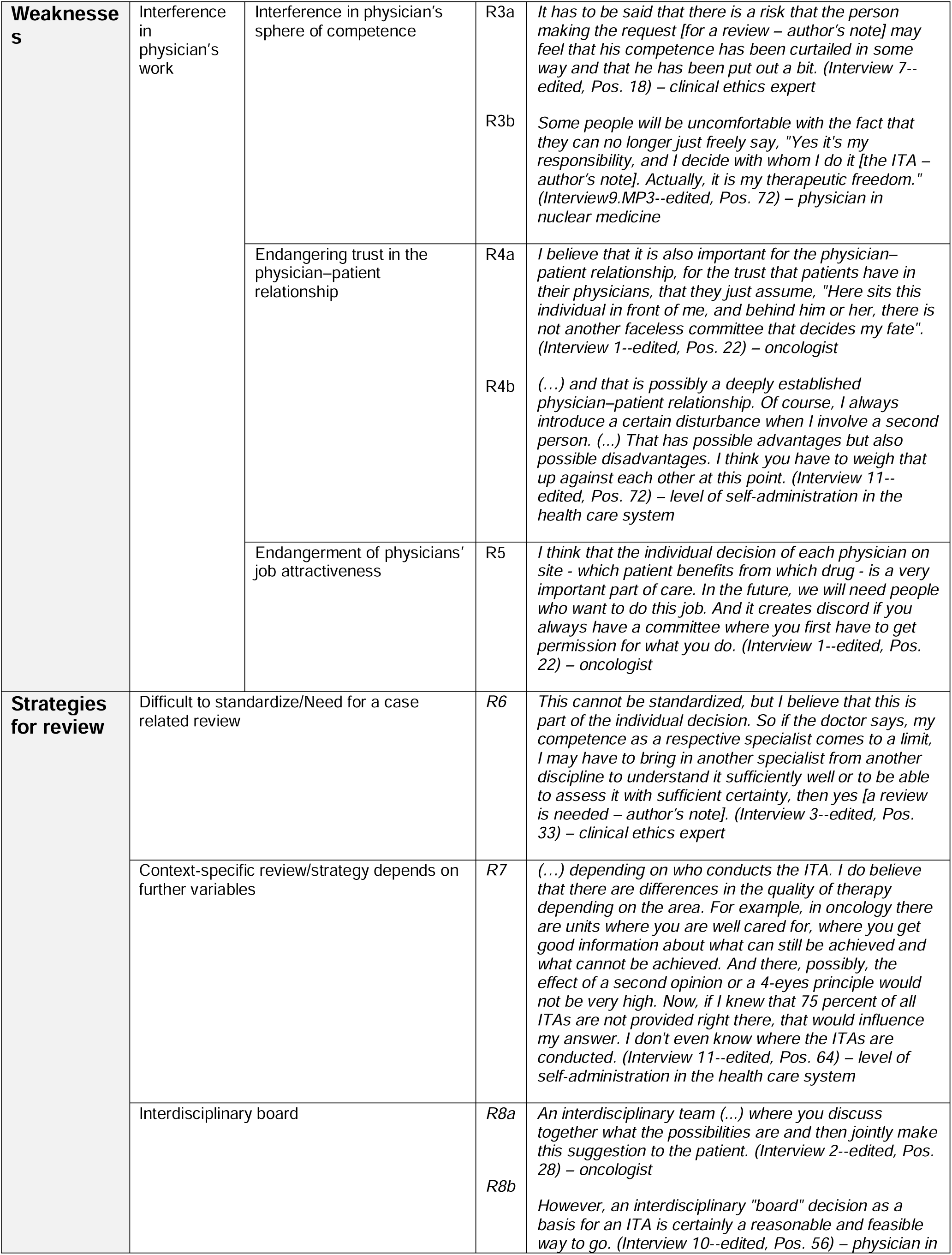
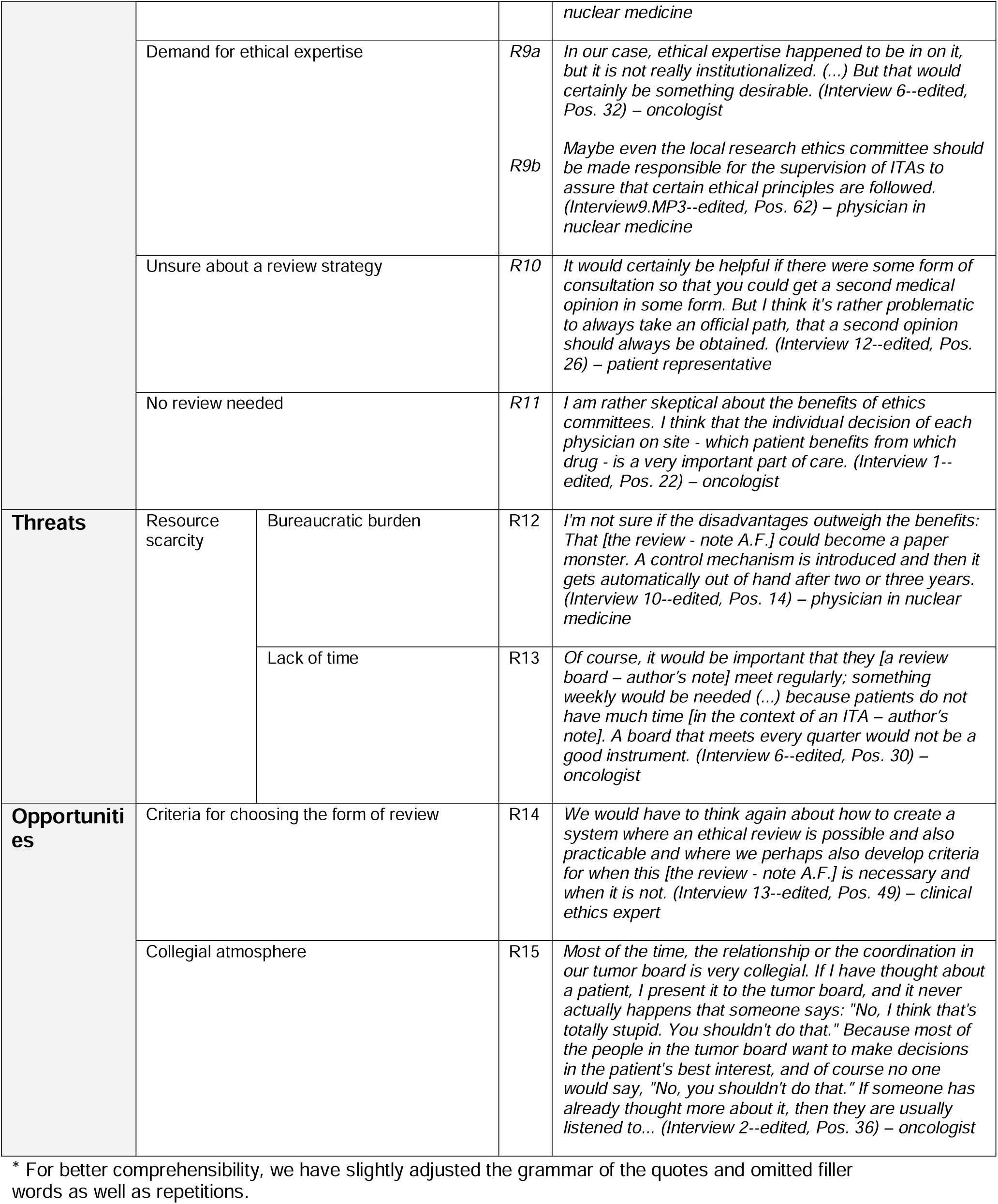
Retrospective and Prospective Evaluation of ITAs

**Table 4*:** Categories of ITAs

| Setting Individual treatment attempts (ITAs) |  |  |  |
| --- | --- | --- | --- |
| Types/Categories | Off-label use | S1a | *The situation in the intensive care unit, I am in the COVID intensive care unit, is that, as expected, nothing is approved for COVID after one year apart from dexamethasone and Clexane. That is, all immunomodulatory drugs that could be applied are administered in the sense of an individual remedy trial. Patients sign a sheet upon admission. In it, we inform the patients that if certain laboratory constellations or certain clinical constellations occur during the course of the disease that suggest an inflammatory syndrome associated with COVID, we would in that case carry out immunomodulation with substances that are not approved for COVID but are approved for various other diseases. (Interview 4--edited, Pos. 10) – physician in intensive care medicine* |
|  |  | S1b | *We have a focus in "city" with minimally invasive epilepsy surgery, and that includes deep brain stimulation and stereotactic laser thermocoagulation. Deep brain stimulation is actually formally approved for one target point, and we have a second one, and we know that it is supposed to be at least as good. There is a constellation of patients for whom we have applied for an ITA more often, either because they were children or people with mental disabilities or because, for example, this target point that we want to reach with deep brain stimulation is no longer available because the patients have brain damage. That's our business, so to speak, that we have to do something like that outside of the formal indication. (Interview8.MP3--edited, Pos. 2) – neurologist* |
|  |  | S1c | *(...) these activities in these programs often result in the discovery of 'targets' that result in 'off-label use'. 'Drug repurposing' plays a large role or is actually the most common outcome of very broad genetic analyses. So, very specifically, a whole genome and RNA analysis in a young patient with an out-treated malignancy, to whom we have nothing to offer in standard therapy, results in a discussion in the molecular tumor board, and there the most frequent 'outcome' is that we recommend a 'repurposing' or a 'repositioning' of an approved substance. (...) that in the context of ITAs, a relatively large amount of immunotherapy is done, certainly also outside the label, especially in younger patients who are still in a good condition, for whom one does not want to give up. (Interview 6--edited, Pos. 2) – oncologist* |
|  | No approval in the EU or in general | S2a | *Basically, it is like this: In oncology, we have above all the setting of chemotherapies that are not yet approved in Germany or in Europe, or rather targeted therapies for certain tumor entities. Either the substance (...) has not yet been approved (...) or we apply to the respective health insurance company for reimbursement of the costs of this drug for a specific patient. In the vast majority of cases, these are seriously ill patients who have already undergone many lines of therapy and where a progression of the disease was nevertheless recorded under the last line. (...) Exactly, so really this individual trial of a drug that is not approved at all, that is exactly in this setting. (Interview 1--edited, Pos. 2) – oncologist* |
|  |  | S2b | *Individual treatment attempts. There are two types and categories. On the one hand, there are the nonapproved, nonestablished procedures (...). (Interview9.MP3--edited, Pos. 12) – physician in nuclear medicine* |
|  | Completely experimental, "from bench to bedside" | S3 | *Now, in the case of completely experimental things, i.e., when you say that someone has a high probability, based on the tumor profile, that you could achieve something with something completely new that is still fresh from the laboratory, which is already being tried at the center once or twice - of course you cannot submit that to the insurance company - and that is then ultimately something that the hospital pays for. (Interview 4--edited,* |
|  |  |  | *Pos. 6) – physician in intensive care/oncologist* |

\* For better comprehensibility, we have slightly adjusted the grammar of the quotes and omitted filler words as well as repetitions.

### Retrospective Evaluation (Monitoring)

#### Strengths

##### System responsibility

Interviewees described several reasons why a basic form of retrospective evaluation of ITAs is an important element of system responsibility. Interviewees, for example, mentioned that a certain form of basic retrospective evaluation of the practice and settings of ITAs would provide important insights that the German health care system currently does not provide (E1). The results of such a basic retrospective evaluation could be used to develop best-practice standards for ITAs that currently do not exist (E2). The systematic documentation of ITAs could further facilitate the sharing of ITA-related experiences with medical peers (E5). An ethicist suggested that the development of standards for retrospective evaluation of ITAs would also become relevant for the evaluation of personalized therapies (E7).

##### Knowledge gain about the intervention used

According to the interviewees, a basic form of retrospective evaluation of ITAs would provide relevant information about the interventions used in an ITA (E8). Some interviewees pointed to the particular interest in gathering data and documenting experiences for ITA interventions used to treat orphan diseases (E8). Some interviewees saw the possibility of a retrospective evaluation as a chance to also make information about negative outcomes of ITAs available for other physicians and scientists. These interviewees complained that, otherwise, only successful ITAs with positive results are published, e.g., as case reports (E10). Furthermore, interviewees suggested that data collected during the conduct of ITAs could serve as a basis to generate hypotheses for future research (E11). Some interviewees mentioned that in rare cases, the results of ITAs could even be used for certain authorization purposes (E12).

#### Weaknesses

##### Validity and practical relevance of evaluation results

Several interviewees were skeptical about the validity and usefulness of retrospective evaluation results (E13-E17). One interviewee mentioned that the data they would gather and then evaluate would be derived from nonideal circumstances in clinical practice and therefore be probably biased in different ways (E15). Another interviewee emphasized the retrospective nature of the data collected in ITAs as a weakness for their usefulness (E14). Interviewees were worried that, in general, it would be difficult to determine which effect to attribute to the drug used in an ITA because, for example, patients treated through ITAs are often treated with several drugs at the same time (E16c). They also pointed out that it might be hard to differentiate between side effects due to the drug or intervention used or a worsening of the patients’ condition due to the natural progression of the often-severe disease (E15b).

While referring to one or more of the above-described challenges, some interviewees questioned whether an intervention-specific retrospective evaluation of ITAs would enable them to draw more general conclusions regarding the risk-benefit ratio of a specific intervention used in an ITA (E16c). Some interviewees went even further and highlighted that because ITAs are so individual, it would be difficult to gather any generalizable information on the effects of ITAs (E16a-b).

##### Difficult distinction between ITAs and research

Some interviewees were critical of the idea of a retrospective evaluation of ITAs because it might be perceived as contradicting the concept of ITAs as a form of therapy and be seen as research instead (E19).

#### Strategies for retrospective evaluation

##### Goals of evaluation

Some clinicians and other stakeholders were interested in an “intervention-specific” retrospective evaluation of ITAs (E22) because this would allow the sharing of experiences with other physicians who had already conducted or planned to conduct similar ITAs (E22b). One interviewee highlighted that evaluating ITAs has become increasingly important for genomic therapies that are specific for individual patients (E22a). Another contrasting idea mentioned by the interviewees was an “intervention-overarching” retrospective evaluation of ITAs. The interviewees in favor of this strategy stressed the importance of determining general facts about ITAs, e.g., in which types of patients ITAs are conducted or how many ITAs are conducted each year. (E21). Such an “intervention-overarching” retrospective evaluation would not assess the effects of a specific intervention used in an ITA but the effects of ITAs as a more general type of intervention/measure in clinical practice.

##### Evaluation method

The interviewees suggested not focusing only on quantitative retrospective evaluations of ITAs but also performing qualitative evaluations to obtain deeper insight into the motivations and circumstances of ITAs (E23). Some interviewees mentioned the possibility of a registry for ITAs (E24), and they had different ideas about the content of the registry, who should complete it and who should have access to it.

#### Opportunities

##### Stakeholders involved in evaluation

The interviewees highlighted that several factors, such as sufficient time (E38), financial allowance (E39), mainly automated documentation (E40), support from specific staff if possible (E42), and certain obligations for documentation, might all contribute positively to a successful retrospective evaluation of ITAs. A participatory approach that involves physicians in the design of retrospective ITA evaluation strategies was further described as a potential facilitator (E36).

##### Structural factors

The interviewees saw it as an opportunity that many patients whom physicians try to treat with ITAs are willing to share their data and are often not scared by potential data security issues (E43).

The legal expert saw no contradiction in retrospectively evaluating ITAs and the concept of an ITA. He argued that retrospectively evaluating the results of an ITA would not change its intention to primarily treat a patient instead of doing research and therefore would not bear the risk of being mistaken for research (E45).

#### Threats

##### Resource scarcity

The interviewees mentioned different hindering factors, such as a lack of time for a retrospective evaluation due to an already high workload of physicians (E28), a lack of money to finance a registry (E27) and other bureaucratic obstacles (E26).

##### Conflicts of interest

Conflicts of interest were described as another potential barrier for the successful implementation of retrospective evaluation. According to the interviewees, conflicts of interest could arise between, e.g., scientists, the pharmaceutical industry and insurance companies (E29). Moreover, the interviewees were worried that physicians could be reluctant to share the results of an ITA or the results of a retrospective evaluation of that ITA because they would want to publish the results on their own to use them for their academic careers (E30).

### Prospective Evaluation (Review)

#### Strengths

##### Good practice for safeguarding favorable risk-benefit ratios

The interviewees pointed out that another expert opinion on whether a certain treatment option with high uncertainty provides a favorable risk-benefit ratio could be seen as a standard element of good practice in ITA decision making (R2).

##### Support of rational risk-benefit assessment

Some interviewees mentioned that a review of risk-benefit judgments can be of particular importance to safeguard rational decision making in situations where physicians who consider ITAs for their severely ill and desperate patients with no other treatment options often feel a strong emotional motivation to offer any possible help (R1).

#### Weaknesses

Some interviewees mentioned that a review, especially an obligatory one, could be seen by physicians as too strong a control of their work and an intervention in their sphere of competence (R9). Another important weakness perceived by some interviewees was a negative influence on the individual physician–patient relationship (R4).

#### Strategies for review implementation

Some interviewees pointed out that the need for and the most suitable format of reviews depend on the context of the ITA (R6-7). This context, however, is often not known because Germany lacks any systematic documentation of ITAs. Other interviewees stressed that the option of obtaining a review by an interdisciplinary board is valuable (R11).

### Categories of ITAs

The comments from the interviewees on the circumstances under which they conduct ITAs and on their overall understanding of ITAs allowed the identification of three categories of ITAs:

- nonstandard off-label use (S1), e.g., drug repurposing in translational oncology (S1c) or for novel diseases in intensive care medicine (S1a);
- interventions that have been tested in at least phase II or phase III trials but that have no authorization in Germany, the European Union or at all (S2);
- highly experimental treatments, from bench to bedside, e.g., new substances that are directly created in the respective laboratory of an oncology center (S3).

## Discussion

We performed 20 in-depth interviews between March and November 2021 to explore stakeholders’ attitudes toward retrospective (monitoring) and prospective (review) evaluations of ITAs in Germany. Neither type of evaluation is currently applied for ITAs in Germany, but these types of evaluations are often applied in similar health care (e.g. organ transplantations, care of premature infants) [14, 15] and research contexts that involve substantial risks, high uncertainty about the likelihood and extent of benefits, and conflicts of interest. The interviewees pointed to several arguments in favor of retrospective evaluation of ITAs, such as the knowledge gain about the setting and circumstances of an ITA and about the types of intervention used. At the same time, the interviewees expressed several concerns regarding the validity and thus the practical relevance of the documentation of ITA cases. The limited validity of documented cases could in turn bias the results of retrospective evaluations. The viewpoints on the prospective review of ITAs addressed several context factors (e.g., medical discipline) that need to be acknowledged in judgments on where review might be more or less needed. They indicated that a review might be necessary for some subgroups of ITAs but that the subgroups are still insufficiently defined. We focus on retrospective evaluation in the discussion because the interviewees discussed this type of evaluation in more depth than prospective evaluation. There was no overall difference in the interviewees’ opinions based on their professional backgrounds.

ITAs involving highly experimental treatments that have not yet been tested in clinical trials might be defined as a subgroup. However, more research is needed to define relevant characteristics for the definition of possible subgroups of ITAs. A systematic documentation of ITAs, such as registry-based documentation, could be piloted for this subgroup. Starting with this subgroup of ITAs also has a strong ethical rationale, as such highly experimental ITAs come with particularly high uncertainty about the risk-benefit ratio [2] and often involve substantial conflicts of interest because those applying the experimental treatments can also be involved academically or even financially in the treatment’s development.

It is likely that most patients are willing to take higher risks in the face of death or severe disability. Furthermore, the determination of what counts as a benefit in such contexts cannot simply be measured in terms of decreased mortality. In the Introduction, we mentioned the example of a highly experimental ITA that was a tremendous success [1]. In another highly experimental ITA, an IgG-based bispecific antibody was given to three patients with metastasized prostate cancer. This led to a reduction in PSA levels (a serum marker for prostate cancer). Whether this surrogate outcome also resulted in patient-relevant outcomes is unknown [27]. We know about these two examples because they were described in scientific publications. Whether these ITA examples are very rare cases in Germany or whether multiple similar cases exist that were not published because of negative outcomes is unclear. Our results indicate that highly experimental ITAs are conducted, but we do not have data about the extent to which they are conducted. Only standardized documentation and retrospective evaluation would allow us to understand the complete picture of benefits and risks resulting from highly experimental ITAs in Germany. However, other scholars have already suggested the concept of a “controlled ITA”, which includes systematic documentation for risky therapies such as somatic gene therapies [28] or xenotransplantation [29].

While several interview participants had positive attitudes toward the documentation of desired and undesired outcomes of ITAs, often the same participants as well as other participants expressed concerns about the validity and practical relevance of the documentation results. The primary goal of ITAs is to treat patients and not to generate generalizable knowledge [30]. The use of innovative therapies (e.g., ITAs) that have not been scientifically validated may lead to an under- or overestimation of their risks and benefits [31]. While the reporting of the context and results of ITAs cannot substitute for clinical trials, the information could serve as a supplement or precursor to scientifically generated data. Furthermore, standardized documentation and the fact that certain ITAs will be evaluated retrospectively could function as an incentive for quality improvement and safety. “Clinical research is one kind of learning activity that might warrant a distinctive kind of oversight, but other kinds of learning are a necessary and commonplace strategy for improving the quality of medical care” [6].

There is a surprising lack of guidance for the conduct of ITAs (including highly experimental ITAs) in the context of the German health care system [32]. This is in contrast to various existing guidelines from professional societies and health care organizations in the USA and Canada for the conduct of innovative practice that offer advice for results reporting and documentation practices [17, 18, 33-35]. Furthermore, a rather theoretical ethics framework for innovative care in pediatrics exists [36]. These guidelines could provide indications for developing guidance for the German concept of ITAs. Without systematically derived knowledge about the current practice of ITAs in Germany, however, it might be difficult to develop valuable and context-specific guidelines. It is likely that there is no one-size-fits-all solution. First-in-human ITAs might need guidance other than nonstandard off-label use.

The interviewees had serious concerns about the feasibility of retrospective evaluation. Piloting systematic documentation and evaluation in a subgroup of ITAs would address this concern to a certain extent. The results of this interview study can inform such piloting activities, further research and further patient and stakeholder activities in this regard.

Similar to retrospective evaluation, the need for prospective evaluation might be more fruitfully discussed for different subgroups of ITAs. Relatively strong normative arguments, for example, can be listed for prospectively reviewing ITAs that employ highly experimental, first-in-human therapies. In these cases, it is particularly difficult for physicians to correctly assess the risks and benefits, and conflicts of interest could be more severe [7]. In the USA, an established review practice for the use of experimental drugs outside of clinical research already exists. Within the Expanded Access Program (for individual patients), a positive vote by a research ethics committee and the U.S. regulatory authority, the Food and Drug Administration (FDA), is required [37]. In the context of the debate around the “Right to Try” law, which was introduced nationwide in the USA in 2018, some authors of the “Compassionate Use & Preapproval Access” working group pointed out that a review by the FDA and a research ethics committee is beneficial for patient safety [37-39].

### Strengths

We are the first to empirically address stakeholders’ attitudes toward the evaluation of ITAs.

### Limitations

Our sample had a gender imbalance, with an overrepresentation of male interviewees. This was partly due to the gender imbalance in certain positions in the German health care system. Our clinical sample consisted only of physicians working at university medical centers. It is possible that ITAs are also performed in other hospitals or on outpatients. We focused on university medical centers because we assumed that most patients for whom standard therapy has failed would be treated in these contexts. It is possible that people who felt a certain need to evaluate the practice of ITAs were more eager to participate in our interviews. The aim of this study, however, was not to quantify the need for practice evaluation. The aim was to describe the qualitative spectrum of arguments and viewpoints illustrating the strengths and weaknesses of such evaluation activities.

## Conclusions

More research on the concrete context and subgroups of ITAs and the close collaboration of stakeholders, such as physicians, professional societies, health care organizations and patients/patient representatives, is necessary to develop a successful evaluation strategy that safeguards the ethically important aspects of the quality and safety of ITAs.

## Supporting information

Supplementary 3 - Interviewguide 1

Supplementary 4 - Interviewguide 2

Supplementary 1 - E-Mail to participants

Supplementary 2 - Study Information and Consent

Supplementary 5 - COREQ Checklist

## Data Availability

The datasets used and analyzed during the current study may be available from the corresponding author on reasonable request. There are legal constraints that prohibit us from making all data publicly available, e.g., they are containing information that could compromise the de-identification of the participating interviewees.

## Acknowledgements

We would like to thank the participating interviewees for their time and willingness to talk to us. We would especially like to acknowledge Tamarinde Haven for providing helpful feedback on the manuscript and on the use of qualitative methods in general. We would like to thank research group Strech for comments on the manuscript prior to submission.

## References

1. Hirsch, T., et al., Regeneration of the entire human epidermis using transgenic stem cells. Nature, 2017. 551(7680): p. 327–332.

2. Hart, D., Heilversuch, in Handbuch Ethik und Recht der Forschung am Menschen. 2014, Springer. p. 47–55.

3. German Federal Supreme Court, Verdict Az. VI ZR 106/13, m.w.N. 2015.

4. Langhof, H. and D. Strech, Off-label use, compassionate use und individuelle Heilversuche: ethische Implikationen zulassungsüberschreitender Arzneimittelanwendungen, in Angewandte Ethik in der Neuromedizin. 2017, Springer. p. 95–105.

5. Heberlein, A., Helfen um jeden Preis?–Historisch fundierte Gründe für das Konzept des „kontrollierten individuellen Heilversuchs “für risikoreiche „individuelle Heilversuche “zur Behandlung einwilligungsunfähiger psychisch kranker Menschen. Ethik in der Medizin, 2013. 25(1): p. 19–31.

6. Earl, J., Innovative Practice, Clinical Research, and the Ethical Advancement of Medicine. The American Journal of Bioethics, 2019. 19(6): p. 7–18.

7. Borysowski, J. and A. Górski, Ethics framework for treatment use of investigational drugs. BMC Medical Ethics, 2020. 21(1): p. 1–10.

8. Caplan, A.L. and A. Ray, The Ethical Challenges of Compassionate Use. JAMA, 2016. 315(10): p. 979–980.

9. Schüklenk, U., Access to unapproved medical interventions in cases of catastrophic illness. Am J Bioeth, 2014. 14(11): p. 20–2.

10. Lenk, C. and G. Duttge, Ethical and legal framework and regulation for off-label use: European perspective. Therapeutics and clinical risk management, 2014. 10: p. 537.

11. Largent, E.A., F.G. Miller, and S.D. Pearson, Going Off-label Without Venturing Off-Course: Evidence and Ethical Off-label Prescribing. Archives of Internal Medicine, 2009. 169(19): p. 1745–1747.

12. Kimmelman, J., Gene transfer and the ethics of first-in-human research: lost in translation. 2010: Cambridge University Press.

13. Rid, A., E.J. Emanuel, and D. Wendler, Evaluating the risks of clinical research. Jama, 2010. 304(13): p. 1472–1479.

14. Institut für Qualitätssicherung und Transparenz im Gesundheitswesen. Qualität der Versorgung sehr kleiner Frühgeborener (NICU). n.d. [cited 2022 21 March]; Available from: https://iqtig.org/qs-verfahren/nicu/.

15. Bundesärztekammer. Richtlinie zur Organtransplantation gemäβ §16 Transplantationsgesetz “Anforderungen an die im Zusammenhang mit einer Organentnahme und -übertragung erforderlichen Maβnahmen zur Qualitätssicherung”. 2001 [cited 2022 21 March]; Available from: https://www.bundesaerztekammer.de/fileadmin/user_upload/downloads/AnfOrga.pdf.

16. World Medical Association Ethics Unit. Declaration of Helsinki. Medical Research Involving Human Subjects. 2013 [cited 2021 15.02.]; Available from: https://www.wma.net/what-we-do/medical-ethics/declaration-of-helsinki/.

17. Stanford University Medical Center. Innovative care guidelines. 2011 [cited 2021 10 December]; Available from: https://stanfordhealthcare.org/content/dam/SHC/healthcare-professionals/medical-staff/policies/innovative-care-guidelines-final-3-18-11copy.pdf.

18. International Society for Stem Cell Research (ISSCR). Guidelines for stem cell research and clinical translation. 2016 [cited 2022 11 January]; Available from: https://www.isscr.org/docs/default-source/all-isscr-guidelines/guidelines-2016/isscr-guidelines-for-stem-cell-research-and-clinicaltranslationd67119731dff6ddbb37cff0000940c19.pdf?sfvrsn=e31478c5_4.

19. Universitätsmedizin Göttingen. Individuelle Heilversuche - Gemeinsames Informationsblatt der Ethik-Kommission für Forschung am Menschen und des Klinischen Ethikkomitees der Universitätsmedizin Göttingen. n.d. [cited x020 25 November]; Available from: https://www.umg.eu/fileadmin/Redaktion/Dachportal/002_Patienten_Besucher/id14_Beratung_Service/id139_Klinisches_Ethikkomitee/Informationsblatt_Individuelle_Heilversuche.pdf.

20. Universität zu Lübeck. Individueller Heilversuch –worauf zu achten ist Gemeinsames Informationsblatt des Klinischen Ethikkomitees des UKSH, Campus Lübecksowie der Ethik-Kommission für Forschung am Menschen der Universität zu Lübeck. n.d. [cited 2020 25 November]; Available from: https://www.uniluebeck.de/fileadmin/uzl_forschung/ethikkommission/SonstigeStudien/Individuelle_Heilversuche_Informationsblatt_Februar2019.pdf.

21. Edwards, R., What is qualitative interviewing? Rosalind Edwards and Janet Holland., in ‘What is?’ research methods series. 2013.

22. Galletta, A., Mastering the semi-structured interview and beyond : from research design to analysis and publication Anne Galletta., in Qualitative studies in psychology. 2013.

23. Philipp, M. and P. Mayring, Qualitative Inhaltsanalyse. 2015: Beltz Verlagsgruppe.

24. Azungah, T., Qualitative research: deductive and inductive approaches to data analysis. Qualitative Research Journal, 2018.

25. Hennink, M.M., B.N. Kaiser, and V.C. Marconi, Code saturation versus meaning saturation: how many interviews are enough? Qualitative health research, 2017. 27(4): p. 591–608.

26. Wieschowski, S., D.S. Silva, and D. Strech, Animal study registries: results from a stakeholder analysis on potential strengths, weaknesses, facilitators, and barriers. PLoS biology, 2016. 14(11): p. e2000391.

27. Zekri, L., et al., An IgG-based bispecific antibody for improved dual targeting in PSMA-positive cancer. EMBO Molecular Medicine, 2021. 13(2): p. e11902.

28. Heinemann, T., et al., Der „kontrollierte individuelle Heilversuch “als neues Instrument bei der klinischen Erstanwendung risikoreicher Therapieformen–Ethische Analyse einer somatischen Gentherapie für das Wiskott-Aldrich-Syndrom. 2006.

29. Heinrichs, B., Forschungsethische Überlegungen zu Humanexperiment und Heilversuch, in Tierische Organe in menschlichen Körpern. 2018, mentis. p. 451–461.

30. Wendler, D., S. Anjum, and P. Williamson, Innovative treatment as a precursor to clinical research. 2021, Am Soc Clin Investig.

31. Bender, S., L. Flicker, and R. Rhodes, Access for the terminally ill to experimental medical innovations: a three-pronged threat. Am J Bioeth, 2007. 7(10): p. 3–6.

32. Borysowski, J., H.J. Ehni, and A. Górski, Ethics codes and use of new and innovative drugs. British journal of clinical pharmacology, 2019. 85(3): p. 501–507.

33. American Medical Association. Ethically Sound Innovation in Medical Practice. n.d. [cited 2022 11 January]; Innovation shares features with both research and patient care, but is distinct from both.]. Available from: https://www.ama-assn.org/delivering-care/ethics/ethically-sound-innovation-medical-practice.

34. Children and Women’s Health Centre of British Columbia. Innovative therapy policy. 2015 [cited 2022 11 January]; Available from: http://policyandorders.cw.bc.ca/resourcegallery/Documents/CW%20Campus%20Wide/Innovative%20Therapy%20Policy%20Draft%20July%2020th%20LPF[5781][6765].pdf

35. American College of Obstetricians and Gynecologists, Innovative practice: ethical guidelines. ACOG Committee Opinion No. 352. Obstetrics and Gynecology 2006. 108(6): p. 1589–1595.

36. Eyadhy, A.A. and S. Razack, The ethics of using innovative therapies in the care of children. Paediatr Child Health, 2008. 13(3): p. 181–4.

37. Chapman, C.R., J. Eckman, and A.S. Bateman-House, Oversight of Right-to-Try and Expanded Access Requests for Off-Trial Access to Investigational Drugs. Ethics & Human Research, 2020. 42(1): p. 2–13.

38. Bateman-House, A. and C.T. Robertson, The Federal Right to Try Act of 2017—A Wrong Turn for Access to Investigational Drugs and the Path Forward. JAMA Internal Medicine, 2018. 178(3): p. 321–322.

39. Folkers, K., C. Chapman, and B. Redman, Federal Right to Try: Where Is It Going? Hastings Center Report, 2019. 49(2): p. 26–36.

