## Supplementary 3 - Interviewguide 1 for "Should the governance of individual treatment attempts (“Individuelle Heilversuche”) include praxis evaluation? Results from qualitative stakeholder interviews"

**Study investigators: Alice Faust, Dr. Lena Woydack, Prof. Dr. Dr. Daniel Strech**
Berlin Institute of Health (BIH)
Translational Research Unit of Charité – Universitätsmedizin Berlin
BIH QUEST Center for Transforming Biomedical Research
Anna-Louisa-Karsch-Str. 2
D-10178 Berlin

### Questions:

1. First, we would like to ask you, in which context do you have contact with individual treatment attempts? For clinicians: Can you describe the circumstances under which you conduct individual treatment attempts. (maybe: How high is the threshold in your team to do an ITA?)

#### Documentation & Data Use

2. How do you think documentation and subsequent data use or evaluation should look like for ITAs? (For clinicians: How do you document ITAs? What do you do with the data you documented, (regardless of them being positive or negative)? Could this be improved?)
3. What would be strengths and weaknesses of such a documentation and data use strategy?
4. Where do you see opportunities and threats for the implementation of such a documentation & data use practice?

#### Transparency

5. The topic of transparency has many facets. What would having an “increased transparency” towards the medical (and scientific) community mean in the context of ITAs?
6. What would be the strengths and weaknesses of an increased transparency?
7. (You suggested…. for gaining an increased transparency.) Where do you see potential threats & opportunities for implementing “an increased transparency strategy”?

#### Review

8. What do you think about a prospective review of ITAs? Which expertise would we need here? What would be important?
9. Which strengths and weaknesses would such a review have? Why?
10. Where do you see opportunities and threats for the implementation of such a review for ITAs?

#### Last part

11. Is there finally something you would like to tell us in this context?
12. Do you know someone, who could be of interest for this study we could approach with an invitation to participate?
