## Supplementary 4 - Interviewguide 2 for "Should the governance of individual treatment attempts (“Individuelle Heilversuche”) include praxis evaluation? Results from qualitative stakeholder interviews"

Berlin, 23.08.21

**Study investigators: Alice Faust, Dr. Lena Woydack, Prof. Dr. Dr. Daniel Strech**
Berlin Institute of Health (BIH)
Translational Research Unit of Charité – Universitätsmedizin Berlin
BIH QUEST Center for Transforming Biomedical Research
Anna-Louisa-Karsch-Str. 2
D-10178 Berlin

### First part

1. First, we would like to ask you, in which context do you have contact with individual treatment attempts? (For clinicians: Can you describe the circumstances under which you conduct individual treatment attempts?)

Currently, individual treatment attempts are subject to a doctor’s freedom of therapy choice with a “special duty of care” (which requires an individual risk-benefit assessment in preparation to individual treatment attempts). In Germany, however, there is no evaluation of the concrete benefits and undesirable effects of individual treatment attempts. We do not know, for example, how many individual treatment attempts are conducted in Germany per year, whether these individual treatment attempts result in the desired effect, and whether they are accompanied by severe side effects.

2. To what extent should there be some form of (overarching or subject-specific) evaluation for individual treatment attempts? *Or:* To what extent is there a need to evaluate the benefits and risks of individual treatment attempts?
    - Why do you think so? / What are the pros and cons of such an evaluation?

### Second part

1. If the interviewees see a need for evaluation: How could this evaluation look like or be implemented into practice?
    - Which threats to a successful implementation could appear?  Which opportunities could support a successful implementation?
2. After we’ve talked about evaluation of individual treatment attempts I would now like to continue with the topic of review. To what extent would patient safety benefit from some form of review in individual treatment attempts? Why is that so?
    - If so, how could such a review look like?

### Last part

1. Is there finally something you would like to tell us in this context?
2. Do you know someone, who could be of interest for this study we could approach with an invitation to participate?
