## Supplementary 1 - E-Mail to participants for "Should the governance of individual treatment attempts (“Individuelle Heilversuche”) include praxis evaluation? Results from qualitative stakeholder interviews"

Dear* …. ,

We are a translational bioethics research group at the QUEST Center, Berlin Institute of Health. We are currently conducting an interview study on "Professional handling of uncertainty in individual treatment attempts (ITAs)". We are interested in your opinion as a ….

In the interviews, we address primarily two topics:

A) **Evaluation**: What are the reasons for or against evaluating the practice of ITAs as a whole?
B) **Review**: What is the case for or against some form of prospective review or second opinion for ITAs?

30-45 minute telephone or video interviews will be conducted. For more details on data protection, please see the attached document. We pay an expense allowance of €150 for interview participation. If you would like to accept this, please sign and return the "Honorarium Form" document, also attached, by post, to the address given in the document. All of your personal information (i.e., name and organizational affiliation) will be pseudonymized and kept strictly confidential. We will publish the results of this interview study in such a way that neither your participation in this interview nor your affiliation with a particular institution will be disclosed.

If you have any further questions, please do not hesitate to contact us.

Kind Regards
Alice Faust (and Lena Woydack and Daniel Strech)
