## Supplementary 2 - Study Information and Consent for "Should the governance of individual treatment attempts (“Individuelle Heilversuche”) include praxis evaluation? Results from qualitative stakeholder interviews"

BIH QUEST
Transforming Biomedical Research

Berlin Institute of Health | Anna-Louisa-Karsch-Straße 2 | 10178 Berlin

**Prof. Dr. Dr. Daniel Strech**
Stellvertretender Direktor QUEST Center

### Professional handling of uncertainty in individual treatment attempts (Individuelle Heilversuche)
### Study information and informed consent

The project addresses the following question: To what extent can precautionary measures contribute to a successful handling of uncertainty in the context of individual treatment attempts? And what SWOTs (strengths, weaknesses, opportunities, threats) are associated with these precautionary measures in the context of individual treatment attempts?

An individual treatment attempt is the use of a (medical) intervention not approved in Germany by a physician on a patient outside of clinical trials for the purpose of his or her therapy.
Individual treatment attempts and clinical research have different primary goals (therapy vs. knowledge gain). However, they also share various characteristics that do not occur in this form in routine care of patients. These include, in particular, a significantly higher degree of uncertainty regarding the potential benefits and risks of the medical intervention in question.

Clinical research is regulated and quality-assured by law and professional practice due to its characteristics, such as high uncertainty and conflicts of interest. Individual treatment attempts fall under the medical freedom of therapy and, despite similar characteristics to clinical research, have no comparable precautionary measures. There is no independent peer review of individual treatment attempts and there is no uniform, systematic documentation of desired and undesired effects. A monitoring of the successes and failures of individual treatment attempts is therefore also not possible.

The introduction of certain precautionary measures in individual treatment attempts is associated with strengths and weaknesses. There would be threats and opportunities for their implementation. We would like to capture these strengths and weaknesses as well as threats and opportunities for implementation objectively and comprehensively in an interview study with relevant stakeholder groups. In total, we plan to conduct about 15-20 semi-structured interviews with experts.

With your expertise, you would make an important contribution to our research.

As a participant, you will be offered an expense allowance of 150 euros.

The interviews are recorded and then transcribed by the staff of a transcription office. This external personnel signs a confidentiality agreement and also receives no information about you or your institution.

In scientific publications, interviews are quoted only in excerpts to ensure that the resulting overall context of text excerpts cannot lead to an identification of your person.

Personal contact data will be stored separately from interview data inaccessible to third parties. After completion of the research project, your contact data will be deleted automatically.

Participation is voluntary and you have the option at any time to withdraw your consent to a recording and transcription of the interview without incurring any disadvantages.

#### Data protection

Your data will be stored and processed in pseudonymized form on Charité servers, in compliance with the EU General Data Protection Regulation (GDPR) and other legislation. Only members of the study team will have access to this data.

After completion of the study, the data will be stored in accordance with the principles of good scientific practice for ten years, after which they will be deleted.
**You have the right to terminate your participation in the study at any time (Art. 21 DS-GVO) and/or to request the deletion of the data held about you (Art. 17 DS-GVO).**

You also have the right to access your personal data stored by the study team (Art. 15 DS-GVO). If you notice that incorrect personal data is stored, you have the right to request a correction (Art. 16 DS-GVO). You also have the right to restrict the analysis of your data (Art. 18 DS-GVO) and/or the transfer of your data (Art. 20 DS-GVO).

**Responsible Data-Controller**: Charité – Universitätsmedizin Berlin, Chariteplatz 1, 10117 Berlin, Germany, Tel: +49 30 450 50, Website: www.charite.de

**Data protection officer:** Questions about the storage and processing of your data, or about your rights to data protection, can be directed to the Charité data protection officer at any time: Datenschutz der Charité (Data Protection Office) – Universitätsmedizin Berlin, Charitéplatz 1, 10117 Berlin, Tel: +49 30 450 580 016,

**Right of appeal** with the Data Protection and Freedom of Information Officer: You have the right to appeal to the competent authority if you have the impression that the processing of your data does not comply with the law: Berliner Beauftragte für Datenschutz und

#### Informed consent

I agree to participate in a semi-structured interview as part of this study (individual treatment attemps). I am aware that my participation is voluntary and that I would not suffer any disadvantage if I did not participate. I am also aware that I can request the deletion of the collected data at any time.

____________________ _____________ ________________________________________________

NAME DATE SIGNATURE
